# “We just considered it [Rift Valley Fever virus] to be over there”: A qualitative study exploring urban perspectives for disease introduction

**DOI:** 10.1101/2022.05.05.22274700

**Authors:** Keli N Gerken, Justinah Maluni, Francis Mutuku, Bryson Ndenga, Caroline Ichura, Luti Mwashee, Makena Mwaniki, Karren Shaita, Stella Orwa, Krish Seetah, A. Desiree LaBeaud

## Abstract

Rift Valley fever virus (RVFV) is a zoonotic arbovirus that is also transmitted to humans from fluids and tissues of infected livestock. Urban areas in Kenya have the hosts, dense vector distributions, and source livestock (often from high-risk locations to meet the demand for animal protein), yet there has never been a documented urban outbreak of RVFV. To understand the likely risk of RVFV introduction in urban communities and guide future initiatives, we conducted Focus Group Discussions with slaughterhouse workers, slaughterhouse animal product traders, and dairy livestock owners in Kisumu City and Ukunda Town in Kenya. For added perspective and data triangulation, in-depth interviews were conducted one-on-one with meat inspector veterinarians from selected slaughterhouses.

Themes on benefits of livestock in the urban setting were highlighted, including business opportunities, social status, and availability of fresh milk in the household. Urban slaughterhouses have formalized meat trading, which, in turn, has resulted in regulation for meat inspection and reduction in local livestock theft. High-risk groups have moderate knowledge about zoonotic diseases and consensus was towards lower personal risk in the urban setting compared to rural areas. Risk assessment was focused on hand hygiene rather than the slaughtering process. There was high reliance on veterinarians to confirm animal health and meat safety, yet veterinarians reported lack access to RVFV diagnostics.

We also highlighted regulatory vulnerabilities relevant to RVFV transmission including corruption in meat certification outside of the slaughterhouse system, and blood collected during slaughter being used for food and medicine. These factors, when compounded by urban vector abundance and dense human and animal populations could create ideal conditions for RVFV to emerge in endemic regions and establish an urban transmission cycle. Here, we present a qualitative study that provides context for urban RVFV introduction risks and insight for adapting current prevention and control measures.

**Author summary:** Rift Valley fever is a globally important zoonotic virus that is also transmitted directly to humans from infected livestock. This qualitative study aimed to explore and understand risk of Rift Valley Fever urban introduction from the perspective of individuals likely at a high-risk of infection in the urban setting. There has never been an urban outbreak of RVFV, however, other arboviruses have caused devastating urban outbreaks driven by urban transmission cycles in vectors. RVFV has a complex epidemiology and urban centers of endemic regions may be particularly vulnerable as they import large quantities of livestock for slaughter and milk to meet the high urban demand for animal sourced foods and have dense vector populations. This qualitative study provides insights on the opinions and lived experiences of urban high-risk groups including slaughterhouse affiliates, livestock owners, and veterinarians. We also demonstrate risks specific to RVFV transmission and regulatory vulnerabilities that would affect ability to detect disease introduction. The results of this study are intended to guide future initiatives aiming to investigate urban introduction of RVFV and determine how the urban disease ecology differs from what has been demonstrated in rural areas.

## Introduction

Rift Valley fever virus (RVFV) is a zoonotic arbovirus with a transmission pathway heavily reliant on socio-cultural interactions with livestock (1). Communities that live in close contact with their livestock are more likely to be exposed to RVFV which has resulted in a confounded understanding of animal contact risk factors (2,3). Other arboviruses have caused devastating outbreaks in urban areas, but there has never been a documented urban outbreak of RVFV (4–6). Limited knowledge of urban RVFV transmission may have resulted in these areas being omitted from mitigation and outbreak planning (7). As the continent of Africa, where RVFV is endemic, is rapidly urbanizing and will soon lead the world in urban growth, a major proportion of future RVFV risk remains unqualified. (8).

The spread of RVFV to new areas primarily relies on movement of infected livestock (9). Urban areas may be particularly vulnerable to RVFV with their dense vector distribution and increased demand for animal-sourced foods (ASFs) and must source livestock and animal products from a wide geographical range. A recent study of two urban areas in Kenya, Kisumu (Western region) and Ukunda (South Coast), revealed a 2% human seroprevalence and highlighted potential sourcing of fresh milk as a potential source of exposure independent of livestock ownership (10). Indeed, the high demand for ASFs in urban areas is well documented in Sub-Saharan Africa, and urban areas often have large tertiary markets (11). To meet demand, animals are imported, alongside a thriving practice in raising livestock within urban contexts (12). Thus, there are two key populations of livestock in urban centers: residential, and regionally imported livestock entering the meat value chain.

In Sub-Saharan African, where RVFV is endemic, recent urbanization over the past 60 years is unprecedented (13). Urban populations in Africa are projected to more than triple over the next four decades and correspond to 21% of the world’s urban population (14). As people migrate to the city for more diverse employment opportunities, better access to healthcare services, and education opportunities, marginalized groups, and the poorest poor face extreme challenges. Inequalities in basic infrastructure result in a lack of piped water, inadequate drainage of sewers and surface water, and higher flooding potential (15). Meanwhile, the lack of proper solid waste management leads to build up of plastic trash that accumulates rainwater and is a preferred breeding site for Aedes spp. mosquitos that transmit arboviruses. (16). Furthermore, and relevant to RVFV transmission, processes of urbanization have occurred rapidly and often over a single generation. Cultural practices may carry different or even higher risk in the urban setting, for example, consumption of raw meat that was slaughtered in a high-volume facility (17). Overall, more preparedness is required for arboviruses because other arboviruses have already proven potential to established urban transmission cycles and have caused devastating outbreaks driven by altered environmental conditions and shifting mosquito populations. (18,19).

In addition to understanding how changing ecological conditions affect risk, the social context of disease is equally important for RVFV. Public health messages that understand and incorporate high-risk populations’ concerns, reality, and perceptions innately have higher success rates for reducing the burden of disease. As highlighted by the recent health messaging during the Covid-19 pandemic, urban populations may be willing yet simply unable to follow health messages particularly when their livelihoods are also at risk (20). For zoonotic livestock diseases, this effect is compounded for those that rely on livestock in urban settings and suddenly lose employment, market access, and purchasing power from closure of livestock markets and restriction of animal movement (21). As RVFV outbreaks have been demonstrated in a variety of climates and ecological conditions (22), understanding the key difference between rural and urban risk factors could allow for inclusive, targeted, public health responses. Similarly, understanding how high-risk groups could be expected to respond can guide population level control measures. To understand urban RVFV risk in greater detail and to guide approaches for developing urban preventive and control measures, we carried out a qualitative study with urban populations we considered to be of highest risk: slaughterhouse affiliates and livestock owners. The data from this study is intended to inform urban preparation, highlight regulatory vulnerabilities for RVFV transmission, and identify opportunities for early detection when regional risk is high.

## Materials and Methods

### Study Design

We carried out a comparative qualitative study between two urban study sites in Kenya in August and September 2021 (Figure 1). A total of ten focus group discussions (FGDs) were conducted to understand the consensus of groups that we considered to be at high-risk for RVFV in urban settings: slaughterhouse affiliates and livestock owners. To triangulate data from each respective slaughterhouse, veterinarians who were meat inspectors responded to in-depth interviews (IDIs) on their perception of risk, experience containing RVFV, and predictions for urban outbreak responses.

**Fig 1:**
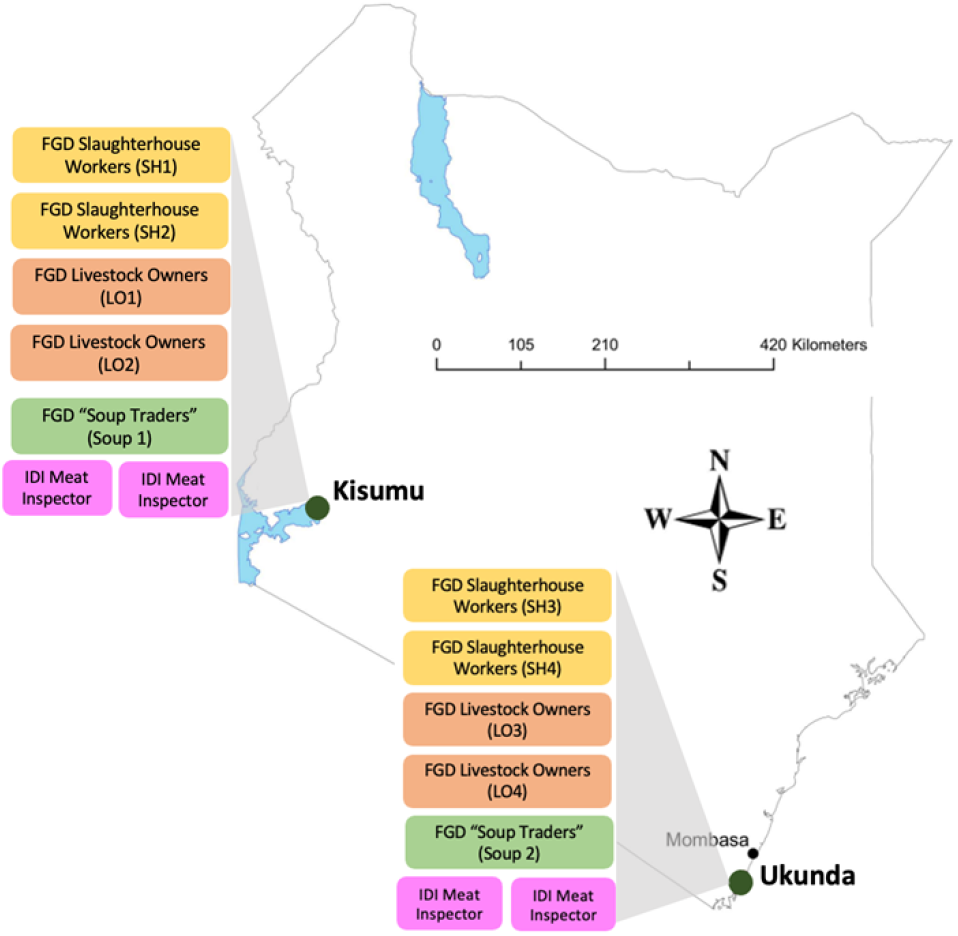
Study design at sites in Kenya. Figure acronyms: LO: Livestock owners; SH: slaughterhouse; FGD: focus group discussion; IDI; in-depth interview Base map: https://africaopendata.org/dataset/kenya-counties-shapefile

### Study Objectives

This study’s main objective was to explore potential pathways of urban RVFV introduction and human exposure from the perspective of those most involved in livestock. Our five specific study objectives are presented in <u>T</u>able 1. Question guides were created to ensure full coverage of the study objectives and homogeneity between the study sites. The guides were piloted with smaller groups (3-5 participants) from adjacent towns to gather baseline information and inform probing techniques for the moderator.

#### Study Areas

The two urban study sites in this study were Kisumu city located on the shores of Lake Victoria in Western Kenya and Ukunda town which is an emerging urban center located on the South coast of Kenya. At both urban sites, nearly all animals are slaughtered in designated slaughterhouse<u>s</u>.

Kisumu (0°5’ 15.2247800 S, 34°46’ 22.328400 E) is the third largest city in Kenya and a business hub at an altitude of 1,100 meters above sea level, and had a population of 610,082 inhabitants in the 2019 census (23). Temperatures are on average 23°C (16-33°C) and total rainfall averages 1,966 mm per year with a relative humidity of 63% (24). The West of Kenya was previously classified as low risk for RVFV outbreaks; however, during the 2018 outbreak, RVFV was detected in adjacent Siaya County of the Western region for the first time (25).

Ukunda (4°16’ 38.8992” S, 39°34’ 9.0012” E) is located on the South coast of Kenya 30 km South of Mombasa at an elevation of 24 meters above sea level with an estimated population of about 100,000 people and represents an area of recent urbanization in Kenya (20). Temperatures are on average 28°C (23-34°C) and total rainfall averages 1,060 mm, per year with a relative humidity of 74% (26). Kwale County has been involved in at least 10 national RVFV outbreaks (27) and recent studies classified Kwale County among the high-risk areas for epizootic RVFV transmission in Kenya (27).

#### Participants and participant recruitment

We worked with key informants and community health volunteers to identify a representative sample of the urban livestock community. We aimed to recruit seven to ten participants for each FGDs involving slaughterhouse workers, slaughterhouse soup traders (described below), and livestock owners. Participants were identified two weeks prior to being invited to participate in the FGDs and attempts were made to capture an equal number of men and women of all age groups and religions.

#### Slaughterhouse workers

Four groups of slaughterhouse workers were recruited from the two main slaughterhouses at each site, and we invited participants that carried out all slaughterhouse activities including the slaughterman, skinners, and meat filleters. The slaughterhouse manager was only included if they participated in the daily activities on the slaughterhouse floor. To be included in this group slaughterhouse employees must have worked at the slaughterhouse for a minimum of six months.

#### Slaughterhouse Soup traders

We identified individuals who purchased various products daily from the slaughterhouse, including the heads, hooves, offal, and blood and invited two groups total, one from each site, to participate in this study. These individuals handle animal products shortly after slaughter and boil the heads and hooves to soup that is sold by the roadside. The most common practice among this group is making soup from these products and therefore, we collectively refer this group as the “slaughterhouse soup traders” (SSTs) throughout this manuscript. They also collect blood to either make local blood sausage (mtura) or sell blood privately to farmers as livestock feed. To be included, SSTs must have purchased either blood or animal products at least three days per week over the past six months.

#### Livestock owners

Four groups of livestock owners, two at each site, were recruited for this study. Ownership in this study was defined as possessing at least one cow, sheep, or goat and we specifically aimed to identify participants involved in local dairy production and business. Although livestock owners with dairy animals were targeted, participants who owned multiple livestock for meat production were also invited.

#### Data collection

The slaughterhouse FGDs took place in a private room or designated area at the respective slaughterhouse. <u>Li</u>vestock owners were invited to meet at a central public location that was identified by key informants and deemed an acceptable travel distance from the participants’ homes. At enrolment, basic demographics were collected for each study participant. FGDs were carried out in Kiswahili except for the SSTs group in Kisumu, which was conducted in the local language, Dhuluo, preferred by all participants. All groups and interviews were audio recorded with a Sony IC audio recorder (ICD-PX470) and transcribed verbatim in English within one week by the moderators and their assistants.

Randomized quality control spot checks were carried out for each transcript by a second transcriber and any discrepancies were discussed.

The first author of this study was present at all the FGDs serving as the note taker and personally led the IDIs. A moderator and assistant moderator were trained from each site and the moderator from the first site (coast) was physically present for the piloting and training at the second site (West) to ensure consistency in data collection. Team training focused on understanding how the study objectives related to RVFV epidemiology and highlighted the importance of minimizing responder bias.

#### Data Analysis

Transcripts were read from which a codebook was developed for thematic analysis. The transcripts were uploaded into NVivo (*Version 1.5, 4577*) and transcripts were coded by two independent coders according to the codebook and organization of codes was discussed in real-time. The coded transcripts were verified by a mutual third coder to ensure consistency.

All FGD data were analyzed together for a collective understanding of urban perspectives and where appropriate, differences between the groups and two study sites were noted. A thematic analysis was carried out to identify prominent themes in the qualitative data. IDIs provided data triangulation from a different perspective and IDI analysis focused on themes identified in the FGDs. Text has been summarized in the results section from detailed ethnographic accounts.

#### Ethics

This study in its entirety was approved by the Institutional Review Board (IRB) at Stanford University (<u>IRB-57869</u>) and the Technical University of Mombasa IRB (TUM ERC EXT/004/2019(R)). Field activities were initiated after receiving a research permit from the Kenyan National Commission for Science, Technology, and Innovation (NACOSTI). Meetings with local administrators (County Commissioners, Deputy County Commissioners, Assistant County Commissioners, Chiefs and Assistant Chiefs) were held to obtain their approvals. Meetings with the community were not conducted due to the then existing Ministry of Health COVID-19 guidelines. Participants were sensitized during recruitment and invited to participate on a voluntary basis. All participants received the same briefing during consenting and were allowed to ask any questions to the group or privately. Before beginning, participants were informed that the study was focused on Rift Valley Fever and their responses could be used to inform policy and disease control in their area. To decrease the risk of responder bias, questions specific to the disease dynamics were deferred to the end of the FGD. The assistant moderator spent a minimum of ten minutes explaining the study procedures and informing participants their rights as a research study participant. Understanding of informed consent was assessed through a checklist at the end of the consent form and each participant signed a consent form and was given a copy. All participants were above Kenya’s adult consenting age of 18 years and were compensated for their lost time and transport costs to attend the meeting point.

## Results

Ninety-one participants from ten different FGDs (n=87) and four IDIs (n=4) were enrolled in this qualitative study and participated until the end of their session. A summary of FGD participant demographics is represented in Table 2. Data saturation was achieved in the final FGDs from the slaughterhouse groups and livestock owners. This was confirmed by codebook developers and no additional themes were added after the final groups. FGD length varied from 46 to 94 minutes. At the coast site (Ukunda), it was challenging to identify female livestock owners despite best efforts from the key informants and snowball sampling techniques. Given the gender ratio for the recruited livestock owners of 13M:4F in Ukunda, it was decided to organize one group of only men and another group with 50% men and 50% women. All slaughterhouse workers at both sites were men; however, women were involved at the slaughterhouse as SSTs. All SSTs in Kisumu were women and the years of experience in this group was comparable to other slaughterhouse staff. The IDIs were with two men and two women and to protect the identity of the meat inspectors, age and location has not been reported.

**Table 1:**
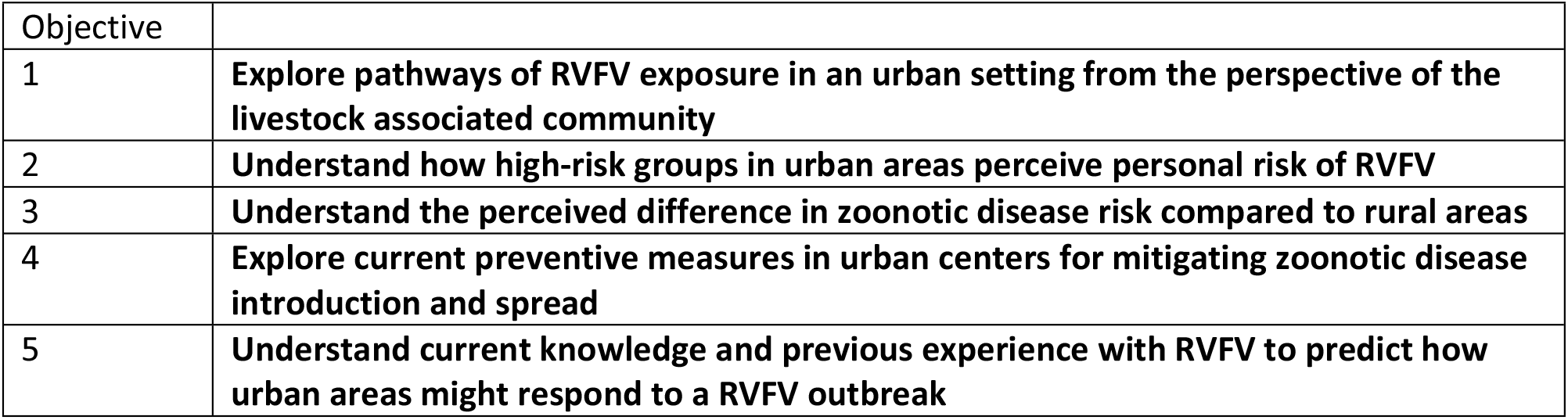
Study Objectives.

| Objective |  |
| --- | --- |
| 1 | **Explore pathways of RVFV exposure in an urban setting from the perspective of the livestock associated community** |
| 2 | **Understand how high-risk groups in urban areas perceive personal risk of RVFV** |
| 3 | **Understand the perceived difference in zoonotic disease risk compared to rural areas** |
| 4 | **Explore current preventive measures in urban centers for mitigating zoonotic disease introduction and spread** |
| 5 | **Understand current knowledge and previous experience with RVFV to predict how urban areas might respond to a RVFV outbreak** |

**Table 2:** Focus Group Discussion Participant Demographics.

|  | Ukunda (coast) |  |  |  |  | Kisumu (West) |  |  |  |  |  |
| --- | --- | --- | --- | --- | --- | --- | --- | --- | --- | --- | --- |
| Demographic | SH1 | SH2 | SST1 | LO1 | LO2 | SH3 | SH4 | SST2 | LO3 | LO4 | Total |
| n=<br>(Gender M/F) | 8 (M) | 9 (M) | 4 (M)<br>6 (F) | 8 (M) | 5 (M)<br>4 (F) | 9 (M) | 8 (M) | 8 (F) | 5 (M)<br>5(F) | 4 (M)<br>4 (F) | 60 (M)<br>27 (F) |
| Median age<br>(years) [range] | 38<br>[32-50] | 37<br>[19-66] | 37<br>[30-62] | 51<br>[26-75] | 48<br>[20-78] | 35<br>[24-49] | 34<br>[26-56] | 52<br>[22-70] | 46<br>[28-69] | 52<br>[22-70] | 41<br>[19-78] |
| Median exp.<br>(years) [range] | 2<br>[10-13] | 14<br>[1-30] | 6<br>[1-10] | 11<br>[2-28] | 10<br>[4-45] | 15<br>[1-30] | 6<br>[3-10] | 9<br>[5-40] | 14<br>[2-52] | 9<br>[5-40] |  |
| Job (SH),<br>Animals Owned<br>(LO), or SH<br>purchases (SST)<br>(n=participants<br>total) | Manger (2)<br>Skinning (6)<br>Intestines (2)<br>Slaughterman (2)<br>Cleaning (2)<br>Filleter (2)<br>Source cattle (1) |  | Heads<br>Blood<br>Hooves<br>Tails<br>Offal | Cos (13)<br>Goats (12)<br>Sheep (4)<br>9 owned<br>multiple species |  | Manager (1) *<br>Skinning (9)<br>Slaughtering (2)<br>Intestines (2)<br>Cleaner (2)<br>*Also skinning |  | <u>All</u><br>Heads<br>Blood<br>(1) | Cows (11)<br>Goats (12)<br>Sheep (1)<br>9 owned<br>multiple species |  |  |
| SH: slaughterhouse; LO: livestock owners; SST: slaughterhouse soup traders<br>M/F: male/female; exp: experience |  |  |  |  |  |  |  |  |  |  |  |

### Socio-cultural dimensions

Analysis focused on themes that were relevant to the study objectives and eight prominent themes (1-8) emerged from the qualitative data. Key messages under each theme have been presented and notable quotations direct from the participants are organized under each theme.

### Theme 1: Social and economic value of urban livestock

Livestock and their products were referred to as urban business opportunities, but they still retained personal sentiment. The sale of animal products paid children’s’ school fees and other daily expenses, while selling livestock was reserved for financial hardships or to offer as dowry for marriage. At least five women in one of the Kisumu livestock owners’ groups and all in the SSTs group regarded that the ability to personally generate income brought them pride. The slaughterhouse workers also expressed pride in providing safe protein to their communities.

The SSTs demonstrated that all animal parts can be used for income generation including the heads, hooves, and blood. Both SSTs groups noted that their products provided customers with a less expensive protein source. Blood that was less fresh could be sold to farmers as feed for pigs (West SSTs) and poultry (all SSTs). All slaughterhouse sales were conducted as private business and the Ukunda slaughterhouse groups highlighted the businessperson who purchased the live animal at market would lose their money if a carcass was condemned.

> *“I have also educated my children through it, so I see it has helped me. My husband had passed on and I was left with them. So, these heads have helped me educate them [my children] …and I am now left the little I get I eat” – Kisumu SST R3*

One major socioeconomic benefit of slaughterhouses mentioned by all four slaughterhouse groups was the reduction in community livestock theft due to the requirement for an official stamp on all marketed meat.

> *<u>“</u>These slaughterhouses have helped in so many ways, cattle theft is very low because when you steal an animal after slaughtering, it isn’t stamped. You’ll have to explain where you got the animal, so even the thieves fear and so it helps the livestock owners too” – Ukunda SH2 R5*

### Theme 2: Connecting human health and animal product consumption

Fresh milk and blood were animal products reported to be linked to human health. Local fresh milk was preferred, particularly for children and babies, because of concerns for chemicals in commercially packed milk. Trust in fresh foreign milk that arrived from surrounding regions was lower than local milk, as they believed the foreign milk had chemicals, adulterants, and water as this was the only way to compete with commercial milk prices.

Among dairy owning livestock owners, it was understood that boiling milk was important for health, but local fresh milk was still sold un-boiled, and it was to be decided by the consumer to boil the milk or not. At least two participants agreed with this and added that some people drink milk direct from the cow and they do not have any health consequences, so therefore, “the dangers are higher than the truth.” However, most participants in the livestock owners’ group recognized milk could cause disease and a commonly known consequence of raw milk consumption was diarrhea. Fresh milk was not just for food but also used as medicine, and tuberculosis (TB), burns, HIV, and arthritis were specifically mentioned to be treated with fresh milk.

Fresh, raw blood was also consumed as a cure for health problems in Kisumu. The slaughterhouse workers shared that blood was collected in plastic jugs and sold to the SSTs who confirmed to purchase these products. The meat inspector veterinarian from this slaughterhouse also affirmed this exchange and recognized that because the blood was pooled from multiple animals it was difficult to remove any if a contributing animal to the pool was condemned. Direct consumption of raw blood from the slaughterhouse was only noted in Kisumu. We did not confirm if the participant’s remark regarding doctors suggesting this practice was in reference to conventional doctors or traditional doctors.

> *“You know blood from an animal is normally warm, then they drink it, the following day they come as well and might go even for a week and then he regains blood quickly that is what the doctor says. He is just someone who has been directed by a doctor that go and drink raw and direct milk from a cow.” – Kisumu SSTs R3*

> *“People believe that that blood boosts their immunity. When they go to the hospital, they are told, especially pregnant mothers, to take blood from the slaughter.” – Vet 3*

### Theme 3: Nutritional demands and livestock movement

Non-local livestock entered urban center for three main reasons: additions to urban herds, arrival at the slaughterhouse to meet the high urban demand for ASFs, or as situational grazers that would arrive and graze for a given time before returning to their origin. Major drivers of livestock movement into urban centers were related to urban nutritional demands of humans and animals. We confirmed animals and animal products must be sourced from distant locations to meet the high demand from the urban population for ASFs. Meanwhile, urban livestock were reported to have limited grazing areas and/or affordable zero-grazing feed options which meant they either had to be grazed along the peri-urban periphery of the city or to scavenge locally on market vegetable waste. Opportunity to mix with local urban animals, either directly or within vector flight range, was noted for slaughterhouse arrivals and situational grazers. Animals that arrive at the slaughterhouse are not slaughtered immediately on arrival and instead holding times were dependent on the orders received by the butcheries. Both Ukunda and Kisumu slaughterhouse workers noted that animals in the holding area never “ran out” as they were always replaced with new arrivals. In Ukunda, holding time was up to one month and the animals primarily walked to the slaughterhouse overnight, even if their origin was more than 200 kilometers away. Animals in holding grazed around the slaughterhouse grounds while they recovered from their transport either on foot or in a lorry. Milk was also a commodity that entered the urban centers in large quantities to meet human nutritional demands. In Kisumu milk was noted to specifically arrive from Eldoret and Nandi. In Ukunda, milk came from nearby Shimba Hills, Lunga <u>L</u>unga, and Tanzania. Milk was reported to quickly sell out as the demand for fresh milk from consumers is very high.

> *“<u>W</u>e first keep them, then slaughter them one by one, depending on the orders we get. We can buy two cows and slaughter on the same day. In short, we keep the cows here for a period of one week to a month depending on the orders.” – Ukunda SH1 R1*

The situational grazers arrived near to the urban sites from far away when there is drought at their home. This practice was more common on the coast of Kenya near to Ukunda when large herds were reported to arrive when they did not have grass available at their respective homes in Kajiado and Garissa counties (two RVFV hotspots in Kenya).

> *“They stay here until rains start, and they call back home and ask if the rains are there or not. <u>I</u>f they are told rains are there, they go back but when they are told the situation is the same, they stay here.” – Ukunda LO2 R4*

> *“I think before December we will receive those visitors. For example, I watched the news yesterday and I found out that in Northeastern its dry to an extent that the cows are dying, so when there is no food, they start coming down here.” – Ukunda LO2 R6*

Urban livestock roaming was a common practice primarily due to feed scarcity. Approximately half of all livestock owners at both sites reported to release their animals daily near to marketplaces for scavenging on vegetable scraps as this was deemed necessary for animals to meet their daily nutritional needs. Two participants noted that despite this, their animals struggled to maintain adequate nutrition. There were additional challenges associated with roaming including being detained by police, poisoning, neighbors abusing animals with knives, dog attacks, snake bites, and consumption of plastic bags. Livestock theft was not noted to be a major concern of this practice.

> *“You can see our cows feeding but they are not strong. The strength we give our cows is by releasing them to struggle through the road to eat people’s kales along the road and to eat garbage that has been pushed together so that they get satisfied.” – Ukunda LO1 R3*

### Theme 4: Veterinarian involvement in slaughter and consumption preferences

Overall, preferences for slaughtering and decisions for meat consumption were highly focused on the presence of the meat inspector veterinarian and his/her post-mortem examination. Trust in the veterinarian was said to overpower any personal opinions and their “final word” was mentioned by all slaughterhouse groups when prompted to discuss food safety. Groups repeatedly assured the moderator that all meat products are inspected, affirming that if the veterinarian had examined a product, then it must be safe for consumption.

The veterinarian’s post-mortem examination, rather than live animal examination, was seen as important as this was the best way to determine if an animal was sick. Mixed responses were received for the best time to call the veterinarian in the case of a suspected sick live animal. If slaughterhouse workers noted any signs of sickness, the animal was either slaughtered for post-mortem examination or the veterinarian was called immediately to examine the live animal. A slaughterhouse worker from Kisumu shared that if an animal arrived at the slaughterhouse and they had been sold it was hit by a car, they preferred to slaughter it quickly to determine if any part of the meat was salvageable. The SSTs who had no direct interaction with the meat inspector, also perceived their products (the heads) to be free of disease since the veterinarian had examined them.

> *“I don’t think there is any risk because the meat is inspected by a doctor. It is confirmed that its healthy, so I don’t feel at any risk”-Ukunda SSTs R2*

> *“When they are bought, we just slaughter them if they have been hit [by a car]. If it was at [nearby rural town], they would have been treated, but here, the medicine is the knife. We involve the vet after we have slaughtered to check the meat.”-Ukunda SH1 R2*

However, animals were sometimes treated for illness before slaughter and all four IDI respondents reported that every urban slaughtered animal receives an antemortem examination the day before, and they were to report to the slaughterhouse for emergencies to determine safety for slaughter if an animal arrival sick or injured. It was also mentioned in Ukunda that if the treatment was with antibiotics, a withdraw period of two weeks was observed.

> *“We have a doctor here who also inspects the animals. He does inspection before we slaughter. So, if it is not okay, he will say that it is supposed to be treated first or it should be returned, it’s not okay. So that also is their work.” -Kisumu SH4 R6*

### Theme 5: Information-focused determinants of risk differential

FGD participants were generally aware that diseases could be transmitted from animals to humans and stated the name of several zoonotic pathogens including anthrax (KSMLO4), bilharzia (KSMLO3), and tetanus (KSM SSTs). The Ukunda SSTs knew they were at risk for zoonotic disease but said that “there is no business which has no risks,” and that even the fire for cooking the products was a health risk for them. The Kisumu SSTs recognized that the slaughterman could become infected with an animal disease by inhaling the “hot blood” from the animal and concluded that while they do have some contact with the blood and animal products, the slaughterhouse workers were touching the animals more, so they were at higher risk. When risk was compared among the slaughterhouse workers groups, both sites identified the slaughterman to be at the highest risk but noted that indeed everyone touches the blood.

The perceived occupational risk of slaughterhouse workers was heavily focused on hygiene which nearly all participants determined to be superior to rural areas because of running water availability. However, one participant in a Kisumu livestock owners group felt that urban populations were at higher risk because of congestion and air pollution.

Slaughterhouse workers could recognize diseases in live animals and explained that examining live animals was important at point of purchase in marketplaces. The animal’s hair and eyes were examined, and other general signs of sickness were related to the animals’ ability to walk, drink water, and eat normally. Post-mortem, slaughterhouse workers could sometimes determine when certain parts of the animal would be condemned even before the vet arrives, but they had to wait for their instruction and oversight. The slaughterhouse workers commonly recognized bruised meat, medication injection sites, parasites in organs and in the intestines, and abnormal livers, knowing these portions would be condemned.

> *“<u>Y</u>ou find the hairs have stood on end. It is difficult to know if it’s the liver that is bad or what, and it’s just there, and later when you slaughter it you find that the liver is bad.” – Kisumu SH3 R1*

> *“There are sick animals at the market, you have to check the health of the animals, some cows can’t even stand, yet we buy and put in the car. When you are at the market it’s hard to know. – Ukunda SH1 R1*

In addition to disease transmission from animals to humans, participants from four out of ten groups connected vectors as a means of infection and explained abundance to be associated with weather and breeding sites. Flies and mosquitoes were said to be more numerous near to places livestock are kept and they were also concerned about tset<u>s</u>e flies and ticks. In Ukunda, one livestock owner participant recognized that if a tick bites another animal and then bites you and your animal that you could get the same disease. Additionally, livestock owners noted that when mosquitoes bite animals they also get malaria. A participant in Kisumu explained that since animals do not sleep under mosquito nets, animals are more at risk of disease transmission from vectors.

> *“Local people can be bitten by mosquitoes; mosquitoes could have bitten an animal then it comes and bit<u>e</u>s you and leaves you with some diseases it got from somewhere.” – Kisumu SH4 R5*

When knowledge sharing was discussed, consensus was reached for wanting more training and demonstrations as fear filled gaps in knowledge. Groups specifically asked to understand transmission pathways, methods of prevention, and disease names so they could determine their risk. Slaughterhouse workers were worried that they touch the blood with their bare hands, but the inspector used a hook. Anthrax was specifically mentioned in Kisumu as one of the biggest fears and the slaughtermen knew not to slaughter if they saw abnormal bleeding in a live animal. One slaughterhouse worker in Kisumu suggested more collaboration between the meat inspect<u>or</u> and the slaughterhouse workers to determine if it was safe to slaughter an animal. The veterinarians reported that they were dedicated to public health and the wellbeing of the slaughterhouse workers, but the slaughterhouse workers had low motivation to learn about the diseases and were focused on business.

> *“I have an opinion, if it’s possible, people like us, make us learn so that we know all the diseases because we just keep cows and only the meat inspector knows the diseases”-Ukunda SH2 R6*

> *“My worry is that, during the process I don’t know what kind of diseases I am exposed to and when it’s a disease that can be transmitted in the blood then we play with blood every day.” – Ukunda SH1 R7*

### Theme 6: Vulnerable regulations

Regulations in place to protect public health were reported to be vulnerable to violations, particularly around meat certification and slaughtering of sick animal<u>s.</u> The slaughterhouse workers group firmly affirmed that it was illegal for any dead animals to enter the slaughterhouse premises. However, at least two participants in the livestock owners group anecdotally described that in recent years, these requirements have been circumvented when meat from sick or dead animals is mis-marketed and mixed in with an officially approved animal. They also stated that these individuals who slaughter sick animals outside of the official system and may have options for receiving fake or corrupt stamps of approval on their meat. One of the main drivers for corruption in meat certification was explained to be the huge financial loss to the owner of the animal. This was noted by the livestock owners’ group in Kisumu rather than by any group at the slaughterhouses.

In the case of an official veterinary meat inspector condemning part or all a carcass, this process has significant oversight by these professionals. In all four IDIs, participants highlighted the importance of strict follow up during the condemnation process and all shared at least one personal story of a community members becoming sick or dying after eating meat that was purchased through a regulatory loophole and therefore, they are strict with follow-up. Notably, one of the meat inspectors from Kisumu revealed that blood is one potential product that can bypass the condemnation process because it is usually pooled early with many other animals and distributed before the inspection occurs.

Vulnerabilities were also present in animal movement restrictions. Data from an Ukunda IDI explained the process for requesting a movement permit and that during high-risk times issuing movement permits was suspended. However, they recognized animals do indeed still move at night undetected or use “short cuts.” In Kisumu, an IDI participant added that while there was an official inspection point at border control with Tanzania, people moved their animals through an unofficial corridor or traveled at night. At the slaughterhouse, movement restrictions were understood to stop disease introduction and they were motivated to comply with regulations because they understood that if the disease arrived, slaughter bans would further affect their livelihoods.

> *“It might not even be the veterinary’s stamp, the butchery people also have their own rubber stamps, and you will find that after the veterinary has stamped theirs on one part of the animal they then stamp on the other pieces or parts and mix with the other pieces.” – Kisumu LO3 R8*

> *“There are those who imagine losing a whole cow, they then make a deal with the slaughter guys, they are then given something little then the cow is taken to the slaughter and slaughtered.” – Kisumu LO3 R10*

### Theme 7: Experience-focused determinants of risk

At least one participant from all FGDs shared an experience with a zoonotic disease control measure, and this contributed to the groups’ self-assessment of risk. At the household level, three livestock owners’ shared practices for reducing mosquitoes around their animals including draining stagnant water and burning branches to create smoke. Livestock owners also shared experience with animal movement restrictions and reactive vaccination.

In urban slaughterhouses, gumboots and a clean laboratory coat were required for work every day and when asked to comment on safety, participants regarded their work uniform to be important in risk reduction. At both sites, the older slaughterhouse workers (>50 years), remembered previously when there were less regulations for slaughtering and believed that the new measures do indeed reduce risk. Slaughterhouse workers also mentioned that their quarterly occupational health checks, included deworming and typhoid vaccinations, are an important component in occupational safety. The slaughterhouse in Kisumu also had some restrictions for animals entering from far away not being able to graze normally with the other animals. At least five slaughterhouse workers recalled animal movement restrictions “nearly closing the meat sector” which greatly impacted their livelihoods. Groups recalled foot and mouth disease outbreaks to be (the most common cause for movement restrictions.

> *“So, you know when they bring the cattle here, we don’t know the problems they have, so when the animals go to graze out there, they mix with the local animals. So, when they mix there, if they have diseases it is transmitted to the others.” – Kisumu SH3 R1*

For RVFV specifically, at least one person in each of the slaughterhouse workers and livestock owners recalled a RVFV outbreak. In Ukunda, the livestock owners noted that during the last outbreak, RVFV was present in the county, but was not seen as an urban threat. One of the meat inspectors confirmed this and expressed frustration for the lack of One Health approaches to RVFV outbreaks, sharing that in the last outbreak, cooperation from the human health sector was low until the first human case was detected in the <u>C</u>ounty Hospital. The majority of FGD participants who knew about RVFV understood it to be a serious disease for livestock and reported that RVFV health messaging focused on avoiding meat and milk consumption, particularly if these products were sourced from the Rift Valley region of Kenya. Most recently, during the 2018 outbreak in Siaya (Western Kenya), three participants in the Kisumu livestock owners group recalled milk that had arrived in the center of town, from the Rift Valley Region, had been poured into the streets after the ban was announced. Another livestock owner in Kisumu, began to fear RVFV when a human in Siaya became infected. An SST in Kisumu also recalled this outbreak and added that everyone feared eating meat at that time. In Kisumu city, at least three livestock owners recalled reactive vaccination of their urban livestock.

A slaughterhouse worker in Kisumu noted that he was never told how to identify an animal that was infected with RVFV over the course of the outbreak, and instead heard about it on the radio. Only one slaughterhouse worker in Ukunda thought he had previously seen signs of RVFV in animals and reported that in 2010 there were numerous deaths in cattle with blood-tinged mucus draining from the nose, though this was never confirmed to be RVFV. For those that had not experienced, RVFV specifically commonly confused it with other fevers and diseases, especially when the other disease has “fever” in the name. East Coast Fever and Yellow Fever were specifically mentioned. Slaughterhouse workers pointed out that when you see something abnormal in the carcass it is challenging to know exactly what disease it is. Other participants didn’t know the disease but attempted to understand the connection with RVFV with the Rift Valley region of Kenya.

> *“Yeah when the announcement came up about RVF, even I can say the milk from Nandi was everywhere here in Kisumu but when the news came up, it’s even hard to get…People just stopped taking the milk so the meat you can’t know because you don’t know where the butchery owner gets their meat from or where they bought it but when it comes to milk many people stopped buying. Most of the people shifted to the processed milk.” – Kisumu LO3 R10*

### Theme 8: Rift Valley Fever human vaccination perceptions

When asked about the potential to be vaccinated against Rift Valley Fever, participants requested more information about the disease and ingredients in the vaccine before they would accept an injection. There was consensus agreement that vaccines worked to prevent diseases and people would be willing to accept a vaccine if they understood their risk. In the slaughterhouse, they were keen to be vaccinated, with some requesting vaccination the same day, because they feared high consequence diseases such as Rift Valley Fever. In Ukunda, livestock owner willingness to be vaccinated was linked to understanding that the livestock community was at higher risk.

> *“It’s a must I know, what will this vaccine do to me, and what is RVF? After knowing what RVF is, I might agree but before that I would not accept the vaccine.” – Kisumu SH3 R7*

> *“So, if you bring that vaccine, do you vaccinate everyone in the homestead or just those who graze or look after the animals because if it can affect me? It can also affect my children and the family. So how you go about it is the most important thing” – Kisumu LO3 R10*

While there was indeed a high acceptance rate, barriers for accepting vaccination were also noted. The main barriers were a lack of a perceived immediate threat, fear of long-term side effects, and confusion with livestock vaccinations. The livestock owners in Ukunda believed that there should be sick animals in their area before they were to accept a vaccination themselves. More hesitancy was present in Ukunda compared to Kisumu as livestock owners were concerned that they would personally receive a cattle injection. A livestock owner in Ukunda said it would be impossible to get people to accept an injection before the disease arrives in their area. In Ukunda, one woman expressed concern the vaccination could cause infertility and all groups wanted to know the potential negative consequences of receiving a vaccine.

> *“Without seeing the disease even if you say you are going to pay me to be vaccinated, still I won’t accept unless the animals here are sick and can transmit to humans” – Ukunda LO1 R1*

> *“Cows, maybe their immunity is higher than that of human beings, so the vaccine that would be administered to cow is it the same as the one that would be administered to me? Are we going to run in the same line as that? The same injection for cows is the same as my injection?” – Kisumu LO4 R4*

## Discussion

This qualitative study has highlighted factors relative to RVFV transmission and key vulnerabilities in urban livestock systems that <u>c</u>ould support successful introduction of RVFV via infected livestock. This risk is intertwined with personal sentiments, challenges faced by the urban livestock community, and private business pressure at the slaughterhouse. Inclusion of the human-centric findings presented here in urban risk management strategies could be expected to allow for earlier and targeted approaches to urban RVFV case identification.

RVFV has been shown to adapt to many different climatic conditions, mammalian hosts, and vectors (22). With the possibility of an imported animal seeding an infection and local herds maintaining or amplifying RVFV, urban disease ecology would differ from rural areas and current measures and surveillance approaches do not account for this. This qualitative study foreshadows how high-risk populations in urban centers could be expected to identify and respond to an introduction. First, for recognizing the first urban livestock cases, slaughterhouses are a key entry point for RVFV as animals enter in high volume from a wide geographical range. Passive syndromic surveillance of livestock alone could be expected to fail in this system as the herd context is required for the most recognizable signs of RVFV: abortion storms and death of young animals. Most animals entering the urban center are adult cattle, which often have inapparent infections (28) and could inadvertently infect slaughterhouse workers, local livestock, or urban mosquitos. The risk to local urban livestock exposure from these imported animals is highlighted by theme three (*Nutritional demands and livestock movement*) as animals have ample opportunity for mixing (either directly or within vector flight range) with non-local animals. If a local animal were to be exposed, the congested living conditions of humans and livestock in the urban setting would deepen the spillover threshold. Notably, lack of feed availability is the root cause of urban livestock roaming, and in the case of an urban outbreak, supporting nutritional demands of these animals could limit unauthorized movement. Moreover, if RVFV is already present in urban vectors, livestock movement bans may be too delayed to have significant effects on the outbreak magnitude and the subsequent threat to livelihoods may not be worthwhile. Risk for urban RVFV viral maintenance is suggested by increased urban abundance of *Aedes*. spp mosquito vectors known to be important in viral amplification phases of a natural outbreak (16), we therefore hypothesize that vector exposures may play a greater role in an urban human exposures. The potential for an undetected introduction coupled with RVFV’s complex epidemiology requires that prevention measures account for all aspects of transmission including human behaviors that affect risk, host interactions with vectors, and opportunities for local animals to be exposed.

Theme two (*Connecting human health and animal product consumption*) of this study demonstrates retention of cultural practices in urban Kenya important in RVFV transmission: consumption of raw blood and fresh milk. Pooled blood purchased directly from slaughterhouses poses a risk to everyone that handles and prepares the product as RVFV is well documented to aerosolize from infected blood, and has been linked to more severe infections (29,30). This exchange of blood from the slaughterhouse also shifts risk into the community where risk factors may be less recognizable, and exacerbates challenges differentiating human RVF disease clinically at health centers from other febrile diseases, including malaria and meningitis (31). Our findings suggest that public health messaging from previous RVFV outbreaks at our study sites to have been primarily focused on consumption of meat and milk, rather than vector exposure, and this is how participants evaluated their risk. This is in contrast to a study conducted in pastoralist communities of Northeastern Kenya where the majority of participants believed RVFV infection to be from mosquito bites rather than from consumption of milk, meat, and blood (32). The authors of that qualitative study reported that while their participants had heard of consumption related risks, their experiences didn’t match their health outcomes and, to them, consumption of meat or milk was not risky for contracting RVFV. In the urban setting, meat and milk consumption is common and there are multiple actors along value chains which can lead to miscommunication about origin and safety of these products. Additionally, there may be implications for blood waste management that contribute to urban transmission dynamics due to the high volume of slaughtered animals and limited physical space because inadequately managed wastewater can breed mosquitos (33). Data on viral presence and stability in blood after slaughter, milk and dairy products, and wastewater from urban slaughterhouses could quantify these public health risks and allow for more specific and directed policy that could be used to build trust and/or be leveraged for surveillance. Furthermore, integrated, and practical approaches to managing these culturally connected risks are required as both livestock owners and SSTs expressed that the ability to purchase and sell their products is important for livelihoods and women’s empowerment.

Our study demonstrated regulatory vulnerabilities in theme six (*Vulnerable regulations*) including selling of unofficially slaughtered meat and unauthorized animal movements that should be considered in urban outbreak investigations and be integrated into future preventive measures. People’s perception of risk impacts their behaviors, and this can either increase the chance of exposure (drinking blood) or be protective (refusing to slaughter dead animals). Therefore, understanding perceptions and challenges that high-risk groups face allows for a more holistic understanding of why regulatory vulnerabilities persist. We highlighted both knowledge-focused and experience-focused factors that affect perceived risk differentials between groups. With these data, we understand current slaughterhouse regulations to be focused on highly prevalent bacterial and parasitic diseases and food safety. This was demonstrated by the participants mentioning hygiene and visual inspection of organs as key measures to determine food safety and reporting that animals are inspected and therefore safe. Reliance on the postmortem examination leads slaughterhouse workers to believe that they must slaughter to determine the ailment of the animal, which neglects their own personal protection from zoonoses. With few individual exceptions, group consensus determined risk to be lower in urban areas compared to rural areas because of improved urban hygiene. Still, fear filled the gaps in participant knowledge, they requested to learn more about their risk of RVFV. Awareness building and education could contribute to better alignment of the knowledge-focused determinants of risk with known RVFV transmission pathways. Messaging would be best focused on the key activities associated with urban slaughterhouse risk and risks in the greater community including handling and consumption of fresh milk and blood, accidental consumption of meat from a sick animal, and urban vector exposure. Awareness building for diseases with alternative transmission mechanisms, such as direct aerosolization of RVFV to humans, can be accomplished in parallel with continued management of other prevalent bacterial and parasitic diseases.

Formalizing meat trade in Kenya by requiring a stamp of inspection from qualified veterinarians has undoubtedly improved key zoonotic diseases and contributed to the improvement of Kenya’s meat safety, although in alignment with other studies, conditions do not always align with Kenya’s Meat Control Act (34). All meat inspector veterinarians in this study shared personal stories about the importance of oversight when condemning a carcass, yet livestock owners reported that corruption and falsification in meat certification does occur, and community-based education of vulnerable groups may alter personal decisions to partake in this risky behavior. For urban introduction of RVFV, slaughterhouse workers and their business partners may be the first line of defense in outbreak control. Full reliance on veterinarians to make key decisions for slaughtering and consumption undermines the ability for slaughterhouse workers to be leveraged in reporting sick animals. In our IDIs, veterinarians understood this public health responsibility, but added that there was insufficient diagnostic support for RVFV, and they fear missing the diagnosis. Underequipping and underpreparing urban meat inspector veterinarians for introduction of RVFV is perhaps the greatest missed opportunity to protect urban public health from RVFV. Furthermore, local One Health initiatives and better administrative cooperation between human health providers and veterinarians are required. If RVFV were to enter an urban center of an endemic region and establish an urban transmission cycle, the human health, and economic effects from loss of livestock would be devastating and concern for re-emergence after would persist every subsequent flooding event. Urban livestock markets would be significantly disrupted and pressure to bypass regulations to maintain livelihoods would be high. Greater understanding of urban transmission risks is warranted now, before the first urban RVFV outbreak.

## Conclusion: Implications for policy to prevent and control urban RVFV

This study has informed understanding of potential pathways of introduction in the context of current practices and urban high-risk groups’ perceived risk. Tracing the pathway of blood, organs, and fresh milk could inform risks in the greater community and allow for preferential testing of RVFV at health center and inform development of inclusive surveillance programs. Possible approaches to prepare urban areas for RVF include education of high-risk groups, community-based awareness building, preventive and reactive livestock vaccination, scenario planning, and surveillance. Passive surveillance is less reliable in urban areas, and we recommend active surveillance, particularly during high-risk times, and development of laboratory procedures for pooled samples to expand cost-effectiveness.

An early and rapid response to RVFV introduction will be required to give urban centers the best opportunity at avoiding infection of urban mosquitos and initiation an urban transmission cycle. Quantifying mosquito vector abundance and diversity at urban slaughterhouses could advise localized vector control strategies. In the meantime, larval source reduction remains important in arboviral disease control, including RVFV (35). Integrating the themes presented here into development of future preventive measures could be expected to be more efficacious and avoid marginalization of vulnerable communities that rely on livestock to support their livelihoods when there is an outbreak. Additionally, a digital record keeping system of urban animal imports could allow veterinarians to carry out local surveillance on suspect animals that enter from high-risk markets known to source animals within high-risk zones. High-risk groups, such as affiliates of the slaughterhouse and livestock owners, of urban areas would likely be the first to identify an urban introduction of RVFV and empowering them to recognize and report suspect cases would assist urban slaughterhouse veterinarians in Kenya that often have overwhelming fast-paced workloads. Urban areas must prepare surveillance frameworks and contingency plans for managing RVFV outbreaks and adapt the context of health messaging from other areas to the perceptions of their own communities.

## Data Availability

The full data has been summarized in the results section of this manuscript.

## Ethical Considerations

This study in its entirety was approved by the Institutional Review Board (IRB) at Stanford University (<u>IRB-5786</u>9) and the Technical University of Mombasa IRB (TUM ERC EXT/004/2019 (R)). The funders had no role in study design, data collection and analysis, decision to publish, or preparation of the manuscript.

## Acknowledgments

This study was funded by NIH Fogarty Global Health Equity Scholars Program NIH, D43TW010540 (Gerken) and NIH, R-01 AI102918 (PI: LaBeaud).

## Author Contributions

**Conceptualization**: Keli Nicole Gerken, Bryson Alberto Ndenga, Francis Maluki Mutuku, Krish Seetah, Angelle Desiree LaBeaud

**Data curation**: Keli Nicole Gerken, Justinah Maluni, Caroline Ichura, Luti Mwashee, Makena Mwaniki, Karren Shaita, Stella Orwa

**Formal analysis:** Keli Nicole Gerken, Justinah Maluni

**Funding acquisition**: Keli Nicole Gerken, Angelle Desiree LaBeaud.

**Investigation**: Keli Nicole Gerken Bryson Alberto Ndenga, Francis Maluki Mutuku, Angelle Desiree LaBeaud.

**Methodology**: Keli Nicole Gerken Bryson Alberto Ndenga, Francis Maluki Mutuku, Luti Mwashee, Krish Seetah, Angelle Desiree LaBeaud.

**Project administration:** Keli Nicole Gerken, Bryson Alberto Ndenga, Francis Maluki Mutuku

**Resources:** Bryson Alberto Ndenga, Francis Maluki Mutuku, Angelle Desiree LaBeaud

**Supervision:** Bryson Alberto Ndenga, Francis Maluki Mutuku, Angelle Desiree LaBeaud

**Validation:** Keli Nicole Gerken, Justinah Maluni, Caroline Ichura

**Visualization:** Keli Nicole Gerken

**Writing – original draft:** Keli Nicole Gerken

**Writing – review & editing**: Keli N Gerken, Justinah Maluni, Francis Mutuku, Bryson Ndenga, Caroline Ichura, Luti Mwashee, Makena Mwaniki, Karren Shaita, Stella Orwa, Krish Seetah, A. Desiree LaBeaud

## Notes

### Competing Interest Statement

The authors have declared no competing interest.

### Funding Statement

This study in its entirety was approved by the Institutional Review Board (IRB) at Stanford University (IRB-57869) and the Technical University of Mombasa IRB (TUM ERC EXT/004/2019 (R)). The funders had no role in study design, data collection and analysis, decision to publish, or preparation of the manuscript.

